# Predictors of adverse outcome in patients with suspected COVID-19 managed in a ‘virtual hospital’ setting: a cohort study

**DOI:** 10.1101/2020.11.09.20228189

**Authors:** Nick A Francis, Beth Stuart, Matthew Knight, Rama Vancheeswaran, Charles Oliver, Merlin Willcox, Andrew Barlow, Michael Moore

**Affiliations:** School of Primary Care, Population Sciences and Medical Education, Faculty of Medicine, University of Southampton, Aldermoor Health Centre, Southampton, SO16 5ST; West Hertfordshire Hospitals NHS Trust, Watford General Hospital, Vicarage Rd, Watford, WD18 0HB

**Keywords:** COVID-19, SARS-CoV2, coronavirus, prognosis, risk factors, community, virtual hospital, cohort

## Abstract

**Objective:** Identify predictors of adverse outcome in a Virtual Hospital (VH) setting for COVID 19.

**Design:** Real-world prospective observational study.

**Setting:** Virtual hospital remote assessment service in West Hertfordshire NHS Trust, UK.

**Participants:** Patients with suspected COVID-19 illness enrolled directly from the community (post-accident and emergency (A&E) or medical intake assessment) or post-inpatient admission.

**Main outcome measure:** Death or (re-)admission to inpatient hospital care over 28 days.

**Results:** 900 patients with a clinical diagnosis of COVID-19 (455 referred from A&E or medical intake and 445 post-inpatient) were included in the analysis. 76 (8.4%) of these experienced an adverse outcome (15 deaths in admitted patients, 3 deaths in patients not admitted, and 58 additional inpatient admissions). Predictors of adverse outcome were increase in age (OR 1.04 [95%CI: 1.02, 1.06] per year of age), history of cancer (OR 2.87 [95%CI: 1.41, 5.82]), history of mental health problems (OR 1.76 [95%CI: 1.02, 3.04]), severely impaired renal function (OR for eGFR <30 = 9.09 [95%CI: 2.01, 41.09]) and having a positive SARS-CoV-2 PCR result (OR 2.0 [95% CI: 1.11, 3.60]).

**Conclusions:** These predictors may help direct intensity of monitoring for patients with suspected or confirmed COVID-19 who are being remotely monitored by primary or secondary care services. Further research is needed to identify the reasons for increased risk of adverse outcome associated with cancer and mental health problems.

**ARTICLE SUMMARY:** *Strengths and limitations of this study:*

- The study uses anonymised data from all patients registered for the virtual hospital between 17/03/20 and 17/05/20, and therefore selection bias is not an issue.
- At the time of this study, this was the only service providing remote follow-up for patients with suspected COVID-19 in the area, and therefore our findings are likely to be relevant to primary care patients receiving remote follow-up.
- We were able to collect reliable data on a wide range of clinical and demographic features, and reliably follow all patients for the primary outcome for at least two weeks following their discharge from the VH.
- We were not able to extract detailed symptom or clinical examination data, and there were significant amounts of missing data for some variables.
- Our study is likely underpowered to detect all predictors, especially in the analysis of our two sub-groups

## BACKGROUND

The COVID-19 pandemic has created unprecedented challenges to healthcare services. Concerns about hospital services being overwhelmed led NHS institutions to develop novel approaches to caring for patients with suspected COVID-19. These include virtual hospitals (VH) where patients who have come to the attention of hospital services and need close monitoring, but do not necessarily need in-patient care, are followed remotely by hospital-based clinicians.^1^ Patients being admitted to such services include those who have presented at accident and emergency (A&E), those referred to the hospital by general practitioners, and those who have had an in-patient admission and are being offered a supported early discharge.

COVID-19 infection is often mild, self-limiting or asymptomatic, but up to 20% of symptomatic individuals may have severe illness.^2^ Identifying those likely to have a worse prognosis is therefore extremely important. Several studies have reported prognostic factors in hospitalised patients, but there have been no studies looking at prognosis in those managed out of hospital via remote patient monitoring services in virtual ward / virtual hospital (VH) settings, who have less severe clinical presentations but may be at risk of deterioration. Factors associated with prognosis are likely to be different in VH patients because they are at a different stage of the disease and/or have less severe symptoms. Understanding factors associated with prognosis in these patients is important in designing services and deciding on admission and escalation criteria, monitoring protocols and discharge criteria. These data are likely to be particularly valuable in informing subsequent waves of COVID-19 and are likely to be relevant to primary care services providing enhanced surveillance of patients with suspected COVID-19 in the community. We therefore set out to identify predictors of adverse outcome in a cohort of patients admitted to a virtual hospital (VH) at one general hospital in England.

## METHODS

This is a prospective observational study using data collected as part of routine clinical care by clinicians working in West Hertfordshire Hospitals. In response to the emerging pandemic, clinicians at Watford General Hospital set up a VH in March 2020. The aim was to reduce pressure on in-patient capacity by providing remote clinical assessment to patients at home in place of hospital admission, or to facilitate early discharge from hospital. Patients with suspected or confirmed COVID-19 were managed in the virtual hospital if they met the inclusion criteria: oxygen saturation >92% on air (or >88% if known to have long-term saturations <92%), resting respiratory rate <20, NEWS < 2, CRP <50, resting HR less than 100, were able to self-isolate and self-care and had access to a telephone or webcam). Patients were triaged into high or low risk pathways for follow-up. Patients were either referred directly from A&E or medical intake (referred to the hospital for assessment but not admitted) (community patients) or were stepped down following a hospital admission (Figure 1).

### Data collection

Participants are patients enrolled in the VH between 17^th^ March and 17^th^ May 2020. Data were recorded as part of routine clinical care with an approved clinical pathway, so participants did not provide informed consent. Data were pseudonymised by staff at West Hertfordshire Hospitals by removing all personal identifying data such as names, date of birth, address. Participants were identified with a unique identifying number, with the key held at West Hertfordshire Hospitals. Pseudonymised data were transferred securely to researchers at the University of Southampton, who analysed the data.

Participants came from one of two routes: 1) patients referred to the VH from A&E or medical intake (community), or 2) patients who were discharged (early) directly to the VH (post-inpatient). At baseline, a general or respiratory consultant working in the VH assessed, examined and investigated patients as part of their clinical care, and documented data in their medical record. Data for this study were subsequently extracted from participants’ medical records. Therefore, data were not collected in a protocolised way but reflect the recording of healthcare data in a busy clinical setting.

Baseline data extracted for the study include: age (calculated from date of birth), gender, smoking status, type of domicile (home, residential home, nursing home, mental health unit, sheltered accommodation, other), comorbid conditions (diabetes, asthma, COPD, other respiratory, cardiovascular disease, chronic kidney disease (CKD), cancer (if recorded in GP or hospital record), connective tissue disorder (CTD), mental health problem), frailty (defined as having a Rockwood score >3 at time of presentation), medications (angiotensin converting enzyme inhibitors (ACEi), angiotensin II receptor blockers (AR2b), non-steroidal anti-inflammatory drugs (NSAIDs), immunosuppressants, oral diabetic medications, insulin, anticoagulants (including direct oral anticoagulants (DOACs)), long-acting beta-agonist (LABA) inhalers, long-acting muscarinic antagonist (LAMA) inhalers, inhaled corticosteroid (ICS) inhalers, beta blockers, proton pump inhibitors (PPI), antidepressants, azithromycin, and hydroxychloroquine), symptoms (presence or absence of: shortness of breath (SOB), cough, fever, chest pain, diarrhoea, headache, myalgia, fatigue). Baseline examination and investigation data extracted for the study include: oxygen saturation, chest x-ray (CXR) result (normal or abnormal), blood tests (white cell count (WCC), lymphocytes, eosinophils, platelets, C-reactive protein (CRP), creatinine, ferritin, D-dimer, troponin). Oxygen saturation levels were categorised as ≤91, 92-93, 94-95, ≥96. Clinicians running the VH attempted to obtain nasal/throat swabs for SARS-CoV-2 testing from all patients. However, during the early phase of the pandemic there was insufficient testing capacity and patients who were not admitted were not tested. SARS-CoV-2 testing was done by PCR at Public Health England (PHE) approved laboratories. Participants were then classified as: COVID-19 positive, negative, inconclusive, or not tested.

Patients referred to the VH were followed up through periodic phone calls to check on their status. High risk patients were followed up by a respiratory consultant on days 2-5, 7, 10, 14 and beyond if needed, whereas lower risk patients were followed up by a consultant physician or GP on days 7 and 14. Decisions about discharge were made by the clinician responsible for the patient based on overall clinical assessment, and were not protocolised. Participants were monitored for two weeks following their initial discharge from the VH, using hospital records to identify overnight re-admission to the hospital and/ or death within this time frame.

### Data analysis

Following data cleaning, standard statistical approaches (proportions, mean and standard deviation) were used to describe the study population, split by route of admission to the VH (from the community or post-inpatient discharge).

Our primary study endpoint was ‘adverse outcome’, defined as death or overnight hospital (re-)admission during the follow-up period (until two weeks after discharge from VH). The relationship between potential baseline predictors and outcome were explored using univariable and then multivariable logistic regression models. Potential predictors included in the model were: gender, age, comorbid conditions, medications, symptoms, oxygen saturation, CXR result, COVID-19 testing, and laboratory test results (WCC, lymphocytes, eosinophils, platelets, CRP, creatinine). All variables were included in a multivariable logistic regression model regardless of the statistical significance of their univariate associations. Backward selection was used with variables retained if p<0.20 (based on log-likelihood). A sensitivity analysis was carried out using a threshold of p<0.10. All adjusted associations are reported as odds ratios with 95% confidence intervals.

We fitted an initial model controlling for the two routes of admission, and we also fitted separate models for these sub-groups.

Multiple imputation using chained equations was used to impute the values of any missing predictors or outcome variables.

### Sample size calculation

Our sample size calculation was based on the minimum required for a multivariable prediction model as set out in Riley et al.^3^ and based on the assumption that 10% of patients experience the outcome and allowed for up to 10 parameters in the final model, with r^2^ of 20% (based on previous literature). Using these parameters and the Stata pmsampsize function,^4^ we calculated a minimum required sample size of 398 patients. Assuming that approximately half of the patients would enter the VH through each of the two routes of admission and allowing for loss to follow-up and missing data, we aimed to include 900 patients.

### Patient involvement

This was an unfunded study set up to analyse existing routinely collected data during a pandemic. Patients were not involved in the design, conduct or reporting of the study.

## RESULTS

Data from the first 900 patients treated in VH were made available for analysis. This included 455 who were admitted directly from the community and 445 who entered the VH post inpatient admissions. Participants were followed for a median of 21 days (range 15 to 46) with very little different between the community (median 21, range 15 to 43 days) and post-inpatient (median 21, range 15 to 46 days) groups. 76 (8.4%) participants experienced an adverse outcome (3 out of hospital deaths, 15 deaths in patients that were (re-)admitted and 58 (re-)admissions to hospital that did not end in death).

The demographic features, comorbid illnesses and current medications of the community and post-inpatient discharge groups, and those who experienced an adverse outcome, are described in Table 1. The population admitted to the VH directly from the community included a greater proportion of females, had a younger average age, more never-smokers and fewer ex-smokers, fewer nursing home residents, fewer patients with physical comorbidities and slightly more with comorbid mental health problems than the post-inpatient group. Baseline symptoms, oxygen saturation levels, and results of investigations are described in Table 2. A slightly larger proportion of the community group reported shortness of breath, cough, chest pain, headache, myalgia and fatigue, than in the post-inpatient group. However, reporting of fever and diarrhoea occurred in a slightly smaller proportion of the community group compared with the post-inpatient group. Normal oxygen saturation levels were much more prevalent in the community group compared with the post-inpatient group (86.5% vs 58.6%) and a smaller proportion of the community group had an abnormal CXR result compared with the post-inpatient group (48.9% vs 77.5%).

**Table 1.** Patient characteristics.

|  | **Post-Inpatient**<br>**(n=445)** | **Community (n=455)** | **Experienced adverse**<br>**outcome (n=76)** |
| --- | --- | --- | --- |
| Experienced adverse outcome | 52/420 (12.4%) | 24/439 (5.5%) | N/A |
| Hospital admission | 49/420 (11.7%) | 24/439 (5.5%) | N/A |
| Death | 16/419 (3.8%) | 2/439 (0.5%) | N/A |
| Female | 202/444 (45.5%) | 275/455 (60.4%) | 37/76 (48.7%) |
| Mean age (s.d.) | 61.0 (17.38) | 48.9 (14.01) | 67.41 (19.88) |
| BAME | 114/438 (26.0%) | 153/448 (34.1%) | 17/76 (22.4%) |
| Smoking |  |  |  |
| - No | 35/190 (18.4%) | 51/163 (31.3%) | 7/39 (18.0%) |
| - Yes | 10/190 (5.3%) | 8/163 (4.9%) | 2/39 (5.1%) |
| - Ex-smoker | 145/190 (76.3%) | 104/163 (63.8%) | 30/39 (76.9%) |
| Domicile |  |  |  |
| - Home | 392/428 (91.6%) | 414/427 (96.7%) | 58/74 (78.4%) |
| - Residential home | 3/428 (0.7%) | 0/427 (0.0%) | 1/74 (1.4%) |
| - Nursing home | 30/428 (7.0%) | 6/427 (1.4%) | 14/74 (18.9%) |
| - Mental unit | 2/428 (0.5%) | 4/427 (0.9%) | 1/74 (1.4%) |
| - Sheltered accommodation | 1/428 (0.2%) | 2/427 (0.5%) | 0 (0.0%) |
| - Other | 0/428 (0.0%) | 1/427 (0.2%) | 0 (0.0%) |
| Comorbid conditions |  |  |  |
| - Diabetes | 110/424 (25.9%) | 52/423 (12.3%) | 27/71 (38.0%) |
| - Frail | 89/430 (20.7%) | 9/426 (2.1%) | 24/74 (32.4%) |
| - Mental health | 133/429 (31.0%) | 142/424 (33.5%) | 31/73 (42.5%) |
| - CKD | 49/426 (11.5%) | 11/424 (2.6%) | 9/73 (12.3%) |
| - CTD | 74/426 (17.4%) | 52/424 (12.3%) | 8/71 (14.2%) |
| - CVD | 44/425 (10.4%) | 13/424 (3.1%) | 9/73 (12.3%) |
| - Cancer | 47/428 (11.0%) | 27/424 (6.4%) | 17/73 (23.3%) |
| - Asthma | 94/431 (21.8%) | 124/427 (29.0%) | 14/74 (18.9%) |
| - COPD | 52/429 (12.1%) | 18/425 (4.2%) | 11/73 (15.1%) |
| - Other respiratory | 30/430 (7.0%) | 17/425 (4.0%) | 4/73 (5.4%) |
| Number of comorbid conditions |  |  |  |
| - None | 100/445 (22.5%) | 159/455 (35.0%) | 13 (17.1) |
| - 1 | 109 (24.5%) | 125 (27.5%) | 16 (21.1%) |
| - 2 | 87 (19.6%) | 97 (21.3%) | 12 (15.8%) |
| - 3 | 58 (13.0%) | 57 (12.5%) | 17 (22.4%) |
| - 4 | 48 (10.8%) | 9 (2.0%) | 7 (9.2%) |
| - 5+ | 43 (9.7%) | 8 (1.8%) | 11 (14.5%) |
| Medications |  |  |  |
| - ACEi | 72/425 (16.9%) | 47/411 (11.4%) | 15/72 (20.8%) |
| - AR2b | 44/424 (10.4%) | 24/410 (5.9%) | 7/72 (9.7%) |
| - Sildenafil | 12/424 (2.8%) | 4/410 (1.0%) | 1/72 (1.4%) |
| - NSAID | 60/425 (14.1%) | 43/410 (10.5%) | 14/72 (19.4%) |
| - Immunosuppressants | 22/425 (5.2%) | 16/410 (3.9%) | 3/72 (4.2%) |
| - LABA | 67/424 (15.8%) | 39/410 (9.5%) | 9/72 (12.5%) |
| - ICS | 85/424 (20.1%) | 68/410 (16.6%) | 12/72 (16.7%) |
| - LAMA | 30/424 (7.1%) | 10/410 (2.4%) | 8/72 (11.1%) |
| - DOAC or other anticoagulant | 77/424 (18.2%) | 22/411 (5.4%) | 19/72 (26.4%) |
| - HQ | 5/424 (1.2%) | 2/410 (0.5%) | 1/72 (1.4%) |
| - Oral diabetes medication | 71/424 (16.8%) | 33/410 (8.1%) | 15/72 (20.8%) |
| - Insulin | 20/424 (4.7%) | 9/410 (2.2%) | 4/72 (5.6%) |
| - Azithromycin | 3/424 (0.7%) | 0/410 (0.0%) | 0 (0.0%) |
| - Beta blockers | 78/425 (18.4%) | 36/410 (8.8%) | 16/72 (22.2%) |
| - PPI | 154/425 (36.2%) | 88/410 (21.5%) | 33/72 (45.8%) |
| - Anti-depressants | 76/424 (17.9%) | 79/410 (19.3%) | 11/72 (15.3%) |
| BAME= black, asian, minority ethnic; CKD= chronic kidney disease; CTD= connective tissue disorder; CVD= cardiovascular disease; COPD= chronic obstructive pulmonary disease; ACEi= angiotensin converting enzyme inhibitor; AR2b= angiotensin II receptor blocker; NSAID= non-steroidal anti-inflammatory drug; LABA= long-acting-beta-agonist; ICS= inhaled corticosteroid; LAMA= long-acting muscarinic antagonist; DOAC= direct oral anticoagulant; HQ= hydroxychloroquine; PPI= proton pump inhibitor. |  |  |  |

**Table 2.**
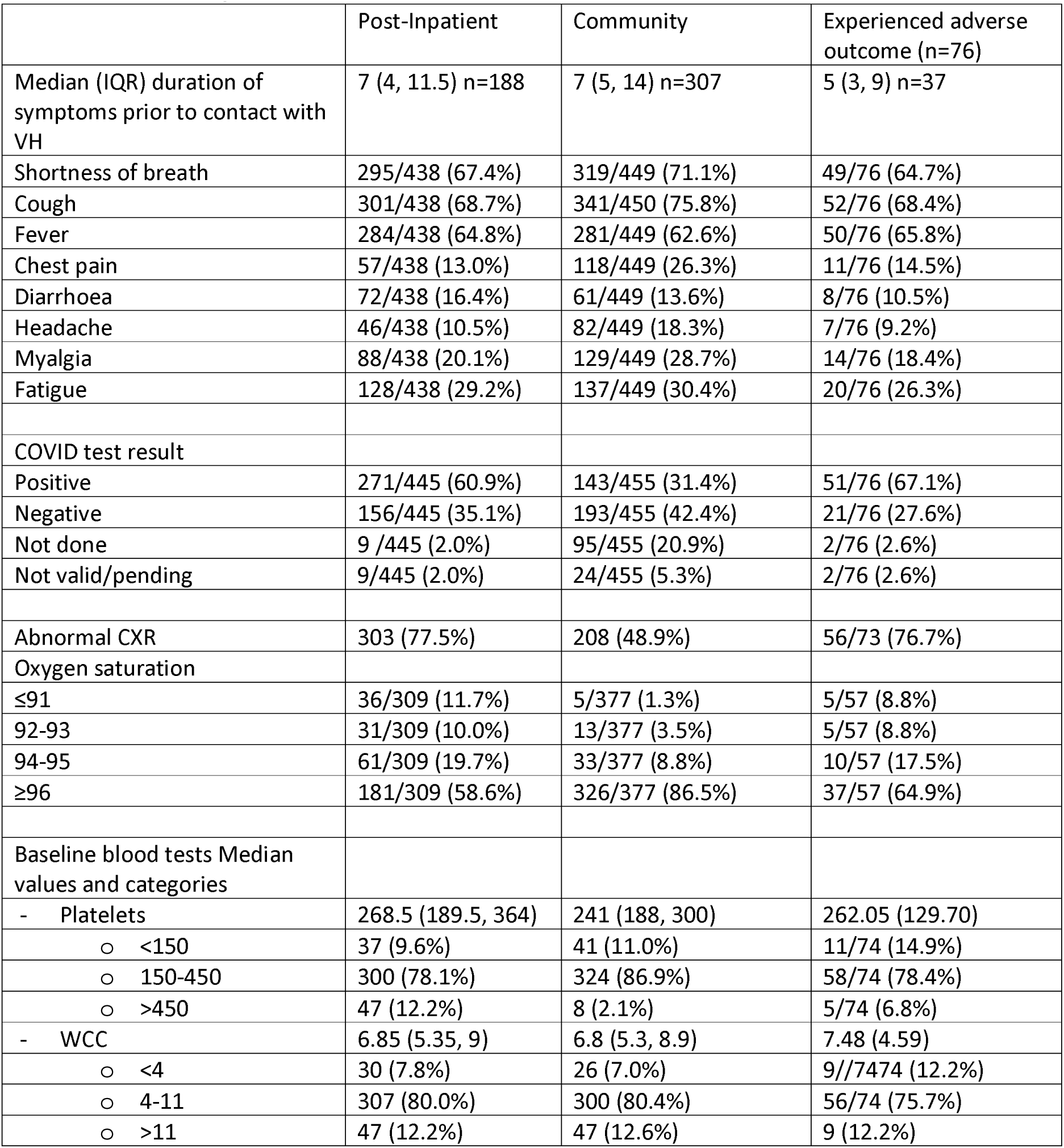
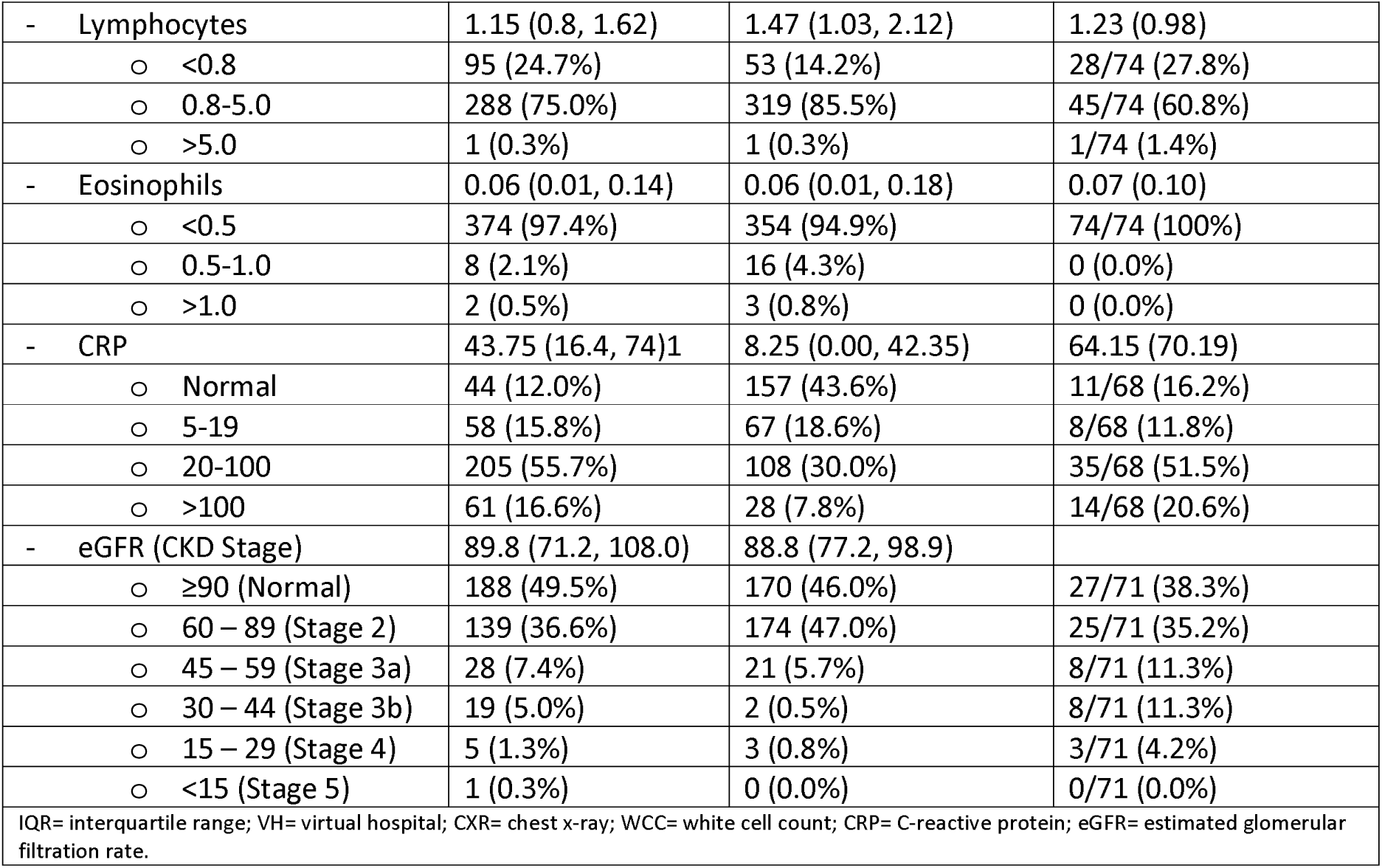
Illness presentation.

763 (84.8%) of the cohort had a valid COVID-19 PCR test result available, with 33 (3.7%) having an invalid test result and 104 (11.6%) not having a test performed (20.9% of the community group and 2.0% of the post-inpatient group). Of those who had a valid test result, 143/336 (42.6%) of the community group had a test that was positive for COVID-19, and 271/427 (63.5%) of the post-inpatient group had a positive test.

### Predictors of adverse outcome

The results of the univariable and multivariable models identifying predictors of adverse outcome in the whole population, controlling for route of admission, are shown in Table 3.

**Table 3.** Association with adverse outcome.

|  | Univariate Odds Ratio (95% CI) | Adjusted Odds Ratio (95% CI) with all variables in the model | Adjusted Odds Ratio (95% CI) retaining only those with p<0.20 (backward selection) |
| --- | --- | --- | --- |
| Community | 0.40 (0.24, 0.66) | 0.67 (0.33, 1.33) |  |
| Male | 1.08 (0.67, 1.74) | 0.56 (0.24, 1.29) |  |
| Age | 1.05 (1.04, 1.07) | 1.04 (1.01, 1.06) | 1.04 (1.02, 1.06) |
| BAME | 0.66 (0.38, 1.16) | 1.05 (0.51, 2.17) |  |
| Comorbid conditions |  |  |  |
| - Diabetes | 2.95 (1.78, 4.89) | 2.31 (1.07, 4.96) | 1.71 (0.95, 3.10) |
| - Mental health | 1.64 (1.00, 2.68) | 1.91 (0.94, 3.89) | 1.76 (1.02, 3.04) |
| - CKD | 2.04 (0.97, 4.29) | 0.33 (0.10, 1.07) | 0.41 (0.15, 1.14) |
| - CTD | 0.82 (0.38, 1.78) | 0.47 (0.16, 1.38) | 0.46 (0.19, 1.09) |
| - CVD | 2.04 (0.96, 4.35) | 1.05 (0.44, 2.41) |  |
| - Cancer | 3.74 (2.03, 6.88) | 3.71 (1.59, 8.64) | 2.87 (1.41, 5.82) |
| - COPD | 2.62 (1.31, 5.23) | 1.58 (0.43, 5.83) |  |
| - Asthma | 0.69 (0.38, 1.25) | 0.76 (0.23, 2.55) |  |
| - Other respiratory | 0.91 (0.35, 2.37) | 0.41 (0.09, 1.82) |  |
| Number of comorbid conditions |  |  |  |
| None | REF | REF |  |
| 1 | 1.45 (0.68, 3.08) | 1.28 (0.53, 3.07) |  |
| 2 | 1.44 (0.64, 3.24) | 0.71 (0.25, 2.03) |  |
| 3 | 3.40 (1.58, 7.30) | 1.10 (0.33, 3.70) |  |
| 4 | 2.99 (1.13, 7.92) | 0.41 (0.10, 1.92) |  |
| 5+ | 5.55 (2.30, 13.33) | 0.78 (0.13, 4.73) |  |
| Medications |  |  |  |
| - ACEI | 1.55 (0.84, 2.80) | 0.96 (0.42, 2.20) |  |
| - AR2b | 1.09 (0.48, 2.50) | 0.64 (0.21, 1.88) |  |
| - Immunosuppressant | 0.83 (0.25, 2.75) | 1.17 (0.52, 2.61) |  |
| - NSAID | 1.79 (0.96, 3.34) | 0.77 (0.19, 3.21) |  |
| - ICS | 1.08 (0.58, 1.99) | 0.91 (0.32, 2.59) |  |
| - DOAC or other anticoagulant | 2.91 (1.62, 5.19) | 1.24 (0.55, 2.80) |  |
| Shortness of breath | 0.80 (0.49, 1.30) | 1.09 (0.58, 2.06) |  |
| Cough | 0.85 (0.51, 1.39) | 1.08 (0.57, 2.07) |  |
| Fever | 1.09 (0.66, 1.78) | 1.29 (0.65, 2.55) |  |
| Chest pain | 0.66 (0.34, 1.27) | 1.36 (0.62, 2.98) |  |
| Diarrhoea | 0.67 (0.32, 1.43) | 0.58 (0.24, 1.39) | 0.55 (0.24, 1.25) |
| Headache | 0.59 (0.26, 1.31) | 1.47 (0.56, 3.88) |  |
| Myalgia | 0.64 (0.35, 1.17) | 0.97 (0.46, 2.02) |  |
| Fatigue | 0.86 (0.51, 1.47) | 0.71 (0.37, 1.35) |  |
| Normal CXR | 0.49 (0.28, 0.86) | 1.10 (0.53, 2.30) |  |
| Oxygen saturation |  |  |  |
| ≤91 | 1.88 (0.72, 4.62) | 0.69 (0.18, 2.63) |  |
| 92-93 | 1.36 (0.49, 3.50) | 0.80 (0.25, 2.62) |  |
| 94-95 | 1.41 (0.69, 2.87) | 0.94 (0.37, 2.38) |  |
| ≥96 | REF | REF |  |
| SARS-CoV-2 test results |  |  |  |
| - Positive | 2.14 (1.27, 3.63) | 1.92 (0.94, 3.93) | 2.00 (1.11, 3.60) |
| - Negative | REF | REF | REF |
| - Invalid/Pending | 0.93 (0.21, 4.15) | 1.02 (0.17, 5.95) | 1.21 (0.24, 5.99) |
| - No swab | 0.28 (0.07, 1.23) | 0.40 (0.08, 2.03) | 0.38 (0.08, 1.76) |
| Platelets |  |  |  |
| - <150 | 1.40 (0.71, 2.79) | 0.96 (0.42, 2.22) |  |
| - 150-450 | REF | REF |  |
| - >450 | 1.03 (0.95, 2.71) | 1.08 (0.35, 3.37) |  |
| WCC |  |  |  |
| - <4 | 1.58 (0.75, 3.33) | 1.42 (0.54, 3.74) |  |
| - 4-11 | REF | REF |  |
| - >11 | 0.95 (0.45, 2.02) | 0.84 (0.33, 2.17) |  |
| Lymocytes |  |  |  |
| - <0.8 | 2.86 (1.72, 4.74) | 1.40 (0.71, 2.74) |  |
| - 0.8-5.0 | REF | REF |  |
| - >5.0 | 13.41 (0.82, 217.82)* | 2.12 (0.01, 341.43) |  |
| Eosinophils |  | 0.14 (0.01, 2.04) |  |
| - <0.5 | REF | REF | REF |
| - 0.5-1.0 | NA | NA | NA |
| - >1.0 | NA<sup>#</sup> | NA | NA |
| CRP |  |  |  |
| - <5 | REF | REF |  |
| - 5-19 | 1.22 (0.49, 3.03) | 0.74 (0.24, 2.28) |  |
| - 20-100 | 2.11 (1.06, 4.19) | 0.94 (0.36, 2.45) |  |
| - >100 | 3.06 (1.33, 7.03) | 1.59 (0.50, 5.09) |  |
| eGFR (CKD Stage) |  |  |  |
| - ≥90 (Normal) | REF | REF | REF |
| - 60 – 89 (Stage 2) | 0.99 (0.55, 1.80) | 1.05 (0.43, 2.54) | 0.78 (0.41, 1.48) |
| - 45 – 59 (Stage 3a) | 1.12 (0.49, 2.53) | 1.52 (0.44, 5.25) | 0.98 (0.41, 2.34) |
| - 30 – 44 (Stage 3b) | 3.90 (1.70, 8.98) | 4.07 (1.04, 16.06) | 2.38 (0.88, 6.46) |
| - <30 (Stage 4/5) | 10.65 (3.38, 33.59) | 22.65 (3.41, 150.70) | 9.09 (2.01, 41.09) |
| * There was only one person in this group.<br># No one in this group had the outcome.<br>BAME= Black, Asian, minority ethnic; CKD= chronic kidney disease; CTD= connective tissue disorder; CVD= cardiovascular disease;<br>COPD= chronic obstructive pulmonary disease; ACEi= angiotensin converting enzyme inhibitor; AR2b= angiotensin II receptor<br>blocker; NSAID= non-steroidal anti-inflammatory drug; ICS= inhaled corticosteroid; DOAC= direct oral anticoagulant; CXR= chest<br>x-ray; WCC= white cell count; CRP= C-reactive protein; eGFR= estimated glomerular filtration rate. |  |  |  |

Univariate analyses found that factors associated with increased odds of adverse outcome were: post-inpatient route of admission; increasing age; comorbid diabetes, COPD, cancer and mental health; anticoagulant medication; abnormal CXR; positive COVID-19 test result; lower lymphocyte count and lower eGFR. The backward stepwise multivariable regression model controlling for route of admission to VH found that factors associated with an increase in the odds of adverse outcome were: increasing age (OR 1.04 [95%CI: 1.02, 1.06] per year), comorbid cancer (OR 2.87 [95%CI: 1.41, 5.82]), comorbid mental health problems (OR 1.76 [95%CI: 1.02, 3.04]), eGFR consistent with CKD Stage 4 or 5 (OR 9.09 [95% CI: 2.01, 41.09] compared with eGFR≥90), and having a positive SARS-CoV-2 PCR result (OR 2.00 [95% CI: 1.11, 3.60] compared with negative test result). The AUROC for the model including these values, after bootstrapping, is 0.76 (95% CI 0.70, 0.83). To shed more light on the results of the regression analyses we reviewed the medical records of participants to further classify the ‘cancer’ and ‘mental health’ comorbid condition categories. This demonstrated that the ‘cancer’ category included cutaneous (20%), breast (20%), haematological (11%), prostate (11%), renal (7%), lung (5%) and other (26%); and the ‘mental health’ category included anxiety (17.9%), depression (29.3%), mixed anxiety and depression (21.7%), alcohol abuse/dependency (6.1%), dementia (8.4%) and other (15.6%).

The results of multivariable models for the community and post-inpatient groups separately are shown in Table 4. In the group referred from the community, only diabetes was found to be a significant predictor of adverse outcome (OR 14.82 [95% CI: 1.14, 192.34]). In the post inpatient group cancer (OR 4.81 [95% CI: 1.42, 16.33]) and eGFR consistent with stage 4 or 5 CKD (OR 34.77 [96% CI: 2.62, 459.77]) were significantly associated with increased odds of adverse outcome and having an ‘other respiratory condition’ was significantly associated with a reduced odds of adverse outcome (OR 0.14 [95% CI: 0.03, 0.76]). The most common conditions coded as ‘other respiratory condition’ were history of: tuberculosis (40%), pulmonary embolism (15%), community acquired pneumonia (9%), asbestosis (6%), sarcoidosis (6%), pulmonary fibrosis (4%), pneumothorax (4%), lung carcinoma (4%).

**Table 4.** Association with adverse outcome in the community and inpatient subgroups.

|  | Inpatient |  |  | Community |  |  |
| --- | --- | --- | --- | --- | --- | --- |
|  | Experienced adverse outcome (n=52) | Univariate Odds Ratio (95% CI) | Adjusted Odds Ratio (95% CI) with all variables in the model | Experienced adverse outcome (n=24) | Univariate Odds Ratio (95% CI) | Adjusted Odds Ratio (95% CI) with all variables in the model |
| Male | 23/52 (44.2%) | 0.65 (0.36, 1.17) | 0.36 (0.13, 0.99) | 14/24 (58.3%) | 2.17 (0.94, 5.00) | 2.36 (0.15, 37.41) |
| Age | 69.8 (21.99) | 1.04 (1.02, 1.06) | 1.03 (1.00, 1.06) | 62.1 (13.17) | 1.07 (1.04, 1.11) | 1.09 (0.98, 1.20) |
| BAME |  | 0.57 (0.27, 1.21) | 0.69 (0.24, 1.97) |  | 1.00 (0.42, 2.42) | 2.49 (0.35, 17.90) |
| Comorbid conditions |  |  |  |  |  |  |
| - Diabetes | 19/51 (37.3%) | 1.86 (1.00, 3.45) | 2.31 (0.85, 6.3) | 8/20 (40.0%) | 4.90 (1.94, 12.37) | 14.82 (1.14, 192.34) |
| - Mental health | 22/52 (42.3%) | 1.84 (1.01, 3.34) | 1.22 (0.47, 3.18) | 9/21 (42.9%) | 1.56 (0.64, 3.79) | 4.55 (0.28, 75.00) |
| - CKD | 7/52 (13.5%) | 1.22 (0.52, 2.87) | 0.30 (0.07, 1.3) | 2/21 (9.5%) | 4.14 (0.84, 20.31) | 0.51 (0.003, 77.02) |
| - CTD | 6/50 (12.0%) | 0.67 (0.27, 1.63) | 0.37 (0.08, 1.6) | 2/21 (9.5%) | 0.89 (0.20, 3.91) | 0.11 (0.002, 5.22) |
| - CVD | 29/51 (56.9%) | 1.56 (0.87, 2.81) | 0.60 (0.18, 1.95) | 10/19 (52.6%) | 2.83 (1.17, 6.87) | 1.78 (0.11, 29.55) |
| - Cancer | 12/52 (23.1%) | 2.67 (1.28, 5.58) | 4.40 (1.34, 14.44) | 5/21 (23.8%) | 5.21 (1.76, 15.40) | 9.94 (0.69, 142.15) |
| - COPD | 9/52 (17.3%) | 1.92 (0.87, 4.21) | 1.22 (0.3, 4.95) | 2/21 (9.5%) | 3.67 (0.86, 15.75) | 8.46 (0.15, 475.34) |
| - Asthma | 11/52 (21.2%) | 0.93 (0.46, 1.89) | 0.94 (0.28, 3.15) | 3/22 (13.6%) | 0.43 (0.13, 1.48) | 0.24 (0.01, 8.03) |
| - Other respiratory | 2/52 (3.9%) | 0.51 (0.15, 1.72) | 0.13 (0.02, 0.95) | 2/21 (9.5%) | 1.86 (0.40, 8.62) | 1.69 (0.04, 72.72) |
| Number of comorbid conditions |  |  |  |  |  |  |
| None | 5/52 (9.6%) | REF | REF | 8/24 (33.3%) | REF | REF |
| 1 | 11/52 (21.2%) | 2.27 (0.78, 6.81) | 2.51 (0.66, 9.61) | 5/24 (20.8%) | 0.79 (0.25, 2.47) | 0.85 (0.10, 7.52) |
| 2 | 9/52 (17.3%) | 2.43 (0.78, 7.54) | 1.46 (0.30, 7.01) | 3/24 (12.5%) | 0.62 (0.16, 2.39) | 0.12 (0.01, 2.27) |
| 3 | 12/52 (23.1%) | 5.24 (1.74, 15.75) | 2.56 (0.43, 15.31) | 5/24 (20.8%) | 1.82 (0.57, 5.81) | 0.52 (0.01, 23.80) |
| 4 | 6/52 (11.5%) | 3.26 (0.96, 11.11) | 1.44 (0.18, 11.76) | 1/24 (4.2%) | 2.31 (0.26, 20.81) | 0.01 (0.00, 13.14) |
| 5+ | 9/52 (17.3%) | 5.31 (1.66, 16.98) | 2.45 (0.20, 30.08) | 2/24 (8.3%) | 6.17 (1.08, 35.53) | 0.19 (0.0002, 167.12) |
| Medications |  |  |  |  |  |  |
| - ACEI | 13/50 (26.0%) | 1.73 (0.87, 3.43) | 1.93 (0.66, 5.66) | 2/22 (9.1%) | 0.71 (0.16, 3.14) | 0.02 (0.0004, 0.81) |
| - AR2b | 4/50 (8.0%) | 0.63 (0.22, 1.85) | 0.76 (0.18, 3.16) | 3/22 (13.6%) | 2.39 (0.66, 8.70) | 0.89 (0.03, 23.24) |
| - Immunosuppressant | 1/50 (2.0%) | 0.28 (0.04, 2.14) | 0.86 (0.27, 2.75) | 2/22 (9.1%) | 2.33 (0.50, 10.92) | 9.80 (0.11, 843.23) |
| - NSAID | 8/50 (16.0%) | 1.23 (0.54, 2.80) | 0.17 (0.02, 1.79) | 6/22 (27.3%) | 3.29 (1.23, 8.81) | 1.73 (0.16, 18.91) |
| - ICS | 11/50 (22.0%) | 1.27 (0.62, 2.57) | 0.84 (0.24, 2.87) | 1/22 (4.6%) | 0.49 (0.10, 2.48) | 1.22 (0.04, 40.98) |
| - DOAC or other anticoagulant | 15/50 (30.0%) | 2.01 (1.03, 3.91) | 1.28 (0.46, 3.58) | 4/22 (18.2%) | 3.87 (1.19, 12.54) | 0.50 (0.02, 10.25) |
| Shortness of breath | 32/52 (61.5%) | 0.76 (0.42, 1.38) | 1.09 (0.48, 2.49) | 17/24 (70.8%) | 1.02 (0.41, 2.53) | 3.66 (0.61, 22.08) |
| Cough | 35/52 (67.3%) | 0.97 (0.52, 1.80) | 1.29 (0.55, 3.07) | 17/24 (70.8%) | 0.77 (0.31, 1.92) | 0.71 (0.12, 4.38) |
| Fever | 31/52 (59.6%) | 0.76 (0.42, 1.37) | 1.14 (0.47, 2.75) | 19/24 (79.2%) | 2.29 (0.84, 6.21) | 3.72 (0.33, 41.95) |
| Chest pain | 8/52 (15.4%) | 1.19 (0.53, 2.68) | 1.99 (0.66, 6.04) | 3/24 (12.5%) | 0.41 (0.12, 1.40) | 0.58 (0.08, 4.44) |
| Diarrhoea | 4/52 (7.7%) | 0.42 (0.15, 1.17) | 0.27 (0.07, 1.00) | 4/24 (16.7%) | 1.28 (0.42, 3.88) | 1.50 (0.18, 12.31) |
| Headache | 3/52 (5.8%) | 0.47 (0.14, 1.56) | 0.74 (0.15, 3.59) | 4/24 (16.7%) | 0.95 (0.31, 2.88) | 4.05 (0.55, 30.08) |
| Myalgia | 7/52 (13.5%) | 0.55 (0.24, 1.26) | 1.05 (0.35, 3.15) | 7/24 (29.2%) | 0.99 (0.40, 2.46) | 0.71 (0.13, 4.01) |
| Fatigue | 12/52 (23.1%) | 0.73 (0.36, 1.45) | 0.56 (0.22, 1.41) | 8/24 (33.3%) | 1.19 (0.49, 2.85) | 0.77 (0.13, 4.61) |
| Normal CXR | 12/49 (24.5%) | 1.09 (0.54, 2.19) | 1.80 (0.62, 5.22) | 5/24 (20.8%) | 0.24 (0.09, 0.65) | 1.00 (0.15, 6.70) |
| Oxygen saturation |  |  |  |  |  |  |
| ≤91 | 4 (7.8%) | 0.94 (0.33, 2.72) | 0.74 (0.17, 3.14) | 1/24 (4.2%) | 5.13 (0.57, 46.24) | 0.96 (0.0003, 2730.82) |
| 92-93 | 4 (7.8%) | 0.82 (0.29, 2.35) | 1.08 (0.28, 4.16) | 1/24 (4.2%) | 2.11 (0.31, 14.49) | 0.54 (0.01, 52.40) |
| 94-95 | 8 (15.4%) | 0.93 (0.39, 2.21) | 0.88 (0.29, 2.71) | 2/24 (8.3%) | 1.59 (0.37, 6.89) | 0.66 (0.03, 14.57) |
| ≥96 | 36 (69.2%) | REF | REF | 20/24 (83.3%) | REF | REF |
| SARS-CoV-2 test results |  |  |  |  |  |  |
| - Positive | 37/52 (71.2%) | 1.67 (0.86, 3.22) | 2.00 (0.78, 5.09) | 14/24 (58.3%) | 2.50 (1.02, 6.15) | 4.08 (0.43, 38.43) |
| - Negative | 13/52 (25.0%) | REF | REF | 8/24 (33.3%) | REF | REF |
| - Invalid/Pending | 1/52 (1.9%) | 1.24 (0.14, 10.63) | 0.41 (0.02, 8.23) | 1/24 (4.2%) | 0.98 (0.12, 8.19) | 2.13 (0.08, 55.79) |
| - No swab | 1/52 (1.9%) | 1.24 (0.14, 10.63) | 5.12 (0.35, 74.63) | 1/24 (4.2%) | 0.24 (0.03, 1.94) | 0.13 (0.01, 3.93) |
| Platelets |  |  |  |  |  |  |
| - <150 | 6/50 (12.0%) | 1.25 (0.49, 3.16) | 0.84 (0.24, 2.73) | 5/24 (20.8%) | 2.00 (0.70, 5.72) | 1.74 (0.23, 13.22) |
| - 150-450 | 40/50 (80.0%) | REF | REF | 18/24 (75.0%) | REF | REF |
| - >450 | 4/50 (8.0%) | 0.64 (0.22, 1.86) | 0.97 (0.25, 3.71) | 1/24 (4.2%) | 1.91 (0.22, 16.19) | 7.47 (0.18, 301.89) |
| WCC |  |  |  |  |  |  |
| - <4 | 8/50 (16.0%) | 2.47 (1.02, 5.98) | 2.21 (0.59, 8.23) | 1/24 (4.2%) | 0.41 (0.05, 3.16) | 0.21 (0.01, 5.81) |
| - 4-11 | 34/50 (68.0%) | REF | REF | 22/24 (91.7%) | REF | REF |
| - >11 | 8/50 (16.0%) | 1.42 (0.61, 3.31) | 1.36 (0.41, 4.52) | 1/24 (4.2%) | 0.27 (0.03, 2.04) | 0.02 (0.0002, 2.40) |
| Lymphocytes |  |  |  |  |  |  |
| - <0.8 | 20/50 (40.0%) | 2.31 (1.24, 4.30) | 1.74 (0.72, 4.25) | 8/24 (33.3%) | 3.37 (1.36, 8.32) | 1.00 (0.11, 9.13) |
| - 0.8-5.0 | 29/50 (58.0%) | REF | REF | 16/24 (66.7%) | REF | REF |
| - >5.0 | 1/50 (2.0%) | N/A | N/A | 0/24 (0.0%) | N/A | N/A |
| Eosinophils |  |  | 0.03 (0.001, 1.15) |  |  | 0.14 (0.01, 1.52) |
| - <0.5 | 50/50 (100.0%) | N/A | N/A | 24/24 (100%) | N/A | N/A |
| - 0.5-1.0 | 0/50 (100.0%) | N/A | N/A | 0/24 (0.0%) | N/A | N/A |
| - >1.0 | 0/50 (100.0%) | N/A | N/A | 0/24 (0.0%) | N/A | N/A |
| CRP |  |  |  |  |  |  |
| - <5 | 6/47 (12.8%) | REF | REF | 5/21 (23.8%) | REF | REF |
| - 5-19 | 5/47 (10.6%) | 0.67 (0.19, 2.34) | 0.52 (0.10, 2.59) | 3/21 (14.3%) | 1.46 (0.34, 6.36) | 0.96 (0.07, 13.06) |
| - 20-100 | 25/47 (53.2%) | 0.96 (0.38, 2.43) | 0.68 (0.17, 2.71) | 10/21 (47.6%) | 2.78 (0.93, 8.30) | 0.65 (0.05, 8.61) |
| - >100 | 11/47 (23.4%) | 1.42 (0.49, 4.13) | 1.43 (0.29, 7.02) | 3/21 (14.3%) | 3.52 (0.79, 15.74) | 1.03 (0.03, 31.33) |
| eGFR (CKD Stage) |  |  |  |  |  |  |
| - ≥90 (Normal) | 20/49 (40.8%) | REF | REF | 7/22 (31.8%) | REF | REF |
| - 60 – 89 (Stage 2) | 15/49 (30.6%) | 0.81 (0.40, 1.63) | 1.15 (0.41, 3.21) | 10/22 (45.5%) | 1.00 (0.30, 3.29) | 0.39 (0.02, 9.63) |
| - 45 – 59 (Stage 3a) | 5/49 (10.2%) | 0.55 (0.17, 1.78) | 0.78 (0.14, 4.27) | 3/22 (13.6%) | 2.62 (0.79, 8.65) | 2.12 (0.07, 61.95) |
| - 30 – 44 (Stage 3b) | 7/49 (14.3%) | 2.25 (0.81, 6.22) | 4.42 (0.73, 26.67) | 1/22 (4.6%) | 7.68 (1.79, 32.81) | 10.37 (0.07, 1616.48) |
| - <30 (Stage 4/5) | 2/49 (4.1%) | 5.69 (1.42, 22.82) | 34.77 (2.62, 459.77) | 1/22 (4.6%) | 24.93 (3.14, 197.81) | 43.36 (0.06, 32422.92) |
| BAME= Black, Asian, minority ethnic; CKD= chronic kidney disease; CTD= connective tissue disorder; CVD= cardiovascular disease; COPD= chronic obstructive pulmonary disease; ACEi= angiotensin converting enzyme inhibitor; AR2b= angiotensin II receptor blocker; NSAID= non-steroidal anti-inflammatory drug; ICS= inhaled corticosteroid; DOAC= direct oral anticoagulant; CXR= chest x-ray; WCC= white cell count; CRP= C-reactive protein; eGFR= estimated glomerular filtration rate. |  |  |  |  |  |  |

## DISCUSSION

### Summary of results

In this observational study of 900 patients admitted to a virtual hospital for remote follow-up of suspected COVID-19 illness, we found that 8.1% of the population were (re-)admitted only 2.0% died during follow-up, giving an overall rate of adverse outcome of 8.4%. Increasing age, comorbid cancer, comorbid mental health, impaired renal function (lower eGFR), and a positive COVID-19 test result were all independently associated with an increased odds of adverse outcome in the combined population, having diabetes was associated with adverse outcome in the community group and history of cancer, eGFR consistent with CKD stage 4 or 5, and not having ‘other respiratory conditions’ were associated with adverse outcome in the post inpatient group.

### Strengths and weaknesses

A strength of this study is the real-world nature of the clinical data used. This was a novel service set up rapidly during a time of crisis, and we included all of the first 900 patients registered with the virtual hospital service. It is reasonably safe to assume that the population included in this study includes the vast majority of those that required monitoring in the community during this period as there were no other services providing remote monitoring of patients that had required a face-to-face assessment in the area at that time. This means that we are unlikely to have the selection bias that characterises many applied research studies. Indeed, by including both patients recruited directly from the community and those who were post-inpatient admission, we have been able to look at predictors in this population suitable for remote follow-up overall, and within each sub-population. For most of the recruitment period there were no general practice hubs assessing patients with suspected COVID-19 in the West Hertfordshire area, and therefore our sample likely includes the majority of patients with suspected COVID-19 that were managed in the community and needed a clinical assessment. A review of the baseline characteristics of these groups demonstrates that we were able to include populations that are likely to be representative of those being followed in the community directly, and those being followed post-inpatient admission. We were able to collect reliable data on a wide range of clinical and demographic features, and reliably follow all patients for the primary outcome for at least two weeks following their discharge from the VH through a review of their hospital records. Another strength is that clinicians were able to validate data collected at baseline during their regular follow-up phone calls. The ‘real world’ nature of our study also poses several limitations. We were not able to extract specific symptom data (such as duration and severity) or data on clinical examination findings (except oxygen saturation) in a consistent and reliable way and there were significant amounts of missing data for some variables (for example oxygen saturation). Because COVID-19 tests were initially only available to inpatients, 20% of the community group did not have a test. We were also unable to collect data on BMI on enough patients to warrant inclusion in our models. It is possible that some patients travelled out of area and were lost to follow-up. However, this seems unlikely given the travel restrictions at the time. Our study is likely underpowered to detect all predictors, especially in the analysis of our two sub-groups. A further weakness of our study is that we did not have a sufficiently large sample to be able to split our sample into development and validation sets. Therefore, our findings need to be validated using an external data set.

### Comparison to other published studies

The most recent version of a ‘living systematic review’ of prediction models for diagnosis and prognosis of covid-19 included 51 studies describing 66 prediction models.^5^ Of the studies included in the review, 32 used data from China, two from Italy, one from Singapore, one from the US, ten international data, two simulated data, and three where the origin of the data was not clear. The majority of the prognostic studies were based on hospitalised patients, and there were no studies of prognosis in virtual hospital settings.

Age has consistently been shown to be a risk factor for poor prognosis in hospitalised,^5-11^ and non-hospitalised ^12^ populations. A large, well conducted study using data from 575 hospitals in China used data from 1590 patients to develop a clinical score for predicting ‘critical illness’ in patients admitted with COVID-19, and validated their score in 710 patients.^11^ Consistent with our findings, they reported age, chest X-ray abnormality, and history of cancer as predictive of adverse outcome. They also identified haemoptysis, dyspnoea, unconsciousness, number of comorbidities, neutrophil to lymphocyte ratio, lactate dehydrogenase (LDH), and direct bilirubin (BR) as predictors. We did not have accurate data on symptoms, and did not have enough data on neutrophil count, LDH or BR to assess these predictors in our model. Another study found that cancer was a risk factor for intubation but not mortality in 5,688 patients admitted to one hospital in New York City with COVID-19.^13^ Although there has been much debate about the effect of COVID-19 on mental health, the association between mental health and adverse outcomes from COVID-19 has not, to the best of our knowledge, been reported in other case series which have been predominantly based around in-patient cohorts. It is possible that those with mental health problems were admitted more frequently because of perceived vulnerability on the part of the clinicians undertaking review assessments, rather than an actual increased risk of physical deterioration. It is also possible that the association between mental health and obesity found in this study was confounded by obesity,^14^ as we were not able to document BMI consistently in this study. However, there may be a variety of reasons why those with mental health disorders are more vulnerable, including reduced levels of activity, impaired socioeconomic status and reduced health care usage for other medical problems. Patients with mental disorders have been noted to have poorer outcomes from other comorbidities, including mortality.^15^ Dementia was a key mental health problem in this cohort, and those with dementia do appear to be at high risk. A study of death certificates in England found that 25.7% of COVID-19 deaths were in patients with dementia, compared to 23.8% of all deaths.^16^ Dementia is clearly associated with other risk factors for poor outcome, but the hypothesis that dementia is associated with a direct causal effect on prognosis warrants further exploration. Close proximity of carers, increased risk of falls, and risk of ‘happy hypoxia’ are possible mechanisms. Other mental illnesses are unlikely to be mentioned on a death certificate, and we have not been able to identify other studies exploring the association between mental health problems and the need for hospital admission for COVID-19.

Given the lack of COVID testing availability during the first few months of the outbreak, diagnostics were only available for patients admitted or those judged to be most at risk. In this cohort, a positive PCR was independently correlated with an increased risk of adverse outcome. This may reflect that testing was initially confined to those patients deemed to be most unwell or may be because patients who did not have COVID-19, or who had a low viral load which was not detected, had a better prognosis.

The inverse association between being coded as having an ‘other respiratory condition’ and experiencing an adverse outcome is difficult to explain. Patients with a history of an assortment of previous and ongoing chronic and acute conditions were lumped together in this category and it is therefore very difficult to interpret the results. Some of the included conditions are associated with immune dysfunction, but it seems unlikely that there is a biological mechanism through which such an assortment of acute and chronic conditions would have a protective effect on adverse outcomes. Therefore, this is more likely to represent a chance finding or unmeasured confounding.

### Implications for policy, practice and research

COVID-19 has changed the face of modern society,^17^ the impact felt from the home to the workplace. The health service has embraced virtual working and remote patient care at scale, in a way never before attempted or achieved. Changes have been rushed through at great pace- and now is the time to reflect, analyse and consider. Same Day Emergency Care (SDEC) (and other out of hospital care pathways) are increasingly being utilised to manage an ever wider range of conditions, ranging from frailty to pneumothorax.^18^ Recent advice from NHS England has advocated the use of oxygen saturation probes in the safe management of COVID-19 as part of remote patient monitoring services.^19^ COVID-19 is a novel disease entity, and unlike many of the other pathologies managed within ambulatory care settings, the natural course of the disease is not yet fully understood. Primary and secondary care practitioners require interim guidance as well as knowledge of clinical practice outside of their own region to guide patient care pending the outcomes of large-scale high-quality research projects.

The relatively low incidence of death and readmission in the multimorbid patients in this study suggest that the clinicians managing this service were able to select and monitor patients in a way that was safe. Comparing outcomes with other approaches to managing these patients, ideally in a randomised trial, would provide more reassurance in this regard.

Our results suggest that in addition to well-known risk factors such as age, clinicians working at the primary-secondary care interface should be aware that patients with coexisting cancer, severely impaired renal function, and mental illness are all at greater risk of hospital admission and/or death, and therefore warrant more careful follow-up. Further research is urgently needed to validate these findings and understand the reasons for the apparent worse prognosis in these patients. There is also a need to assess the most cost-effective approaches to monitoring and supporting patients in the community with suspected/confirmed COVID-19 who do not (yet) require hospital admission.

### Conclusions

This observational study of a real-world remote monitoring VH service, set up rapidly during the onset of the worst pandemic seen in decades, has demonstrated that it was possible to set up a service that resulted in a low incidence of deaths (2.0%) and readmissions (8.1%). When planning and commissioning services in primary and secondary care to manage patients with COVID-19 during this ongoing pandemic, we would suggest that the risk factors for deterioration identified in this cohort, namely age, significant renal impairment (CKD stage 4-5), history of cancer, and history of mental health problems, should merit more intensive follow up and monitoring.

## Data Availability

The data that support the findings of this study are available from West Hertfordshire Hospitals NHS Trust but restrictions apply to the availability of these data, which were used under license for the current study, and so are not publicly available. Data are however available from the authors upon reasonable request and with permission of West Hertfordshire Hospitals NHS Trust.

## FUNDING

This research received no specific grant funding from any funding agency in the public, commercial or not-for-profit sectors.

## ETHICS APPROVAL AND CONSENT TO PARTICIPATE

Regulatory and ethical approval for the study were provided by the Health Research Authority and Chelsea Research Ethics Committee (REC reference: 20/HRA/2342). The data used in this study were collected as part of routine healthcare during a pandemic. Participants did not provide consent to participate. Data were extracted from medical records by clinicians providing care for the patients and anonymised data were provided to the research team at the University of Southampton. No identifiable data left the hospital.

## COMPETING INTERESTS

The authors declare that they have no competing interests.

## AUTHOR CONTRIBUTIONS

All authors contributed to the conception and design of the study. MK, RV, CO and AB contributed to data collection. BS led the data analysis of the data. All authors contributed to data interpretation. NF wrote the first draft of the paper and all authors contributed to revising the manuscript.

## ACKNOWLEDGEMENTS

We would like to acknowledge Dr David Evans and Mr Alex Newland Smith who provided a lot of assistance in collecting the data.

## Notes

### Competing Interest Statement

The authors have declared no competing interest.

### Clinical Trial

This study uses medical record data and therefore was not registered

### Summary of Updates

There was an error in the abstract with an odds ratio being replaced with the letters, "OR Mike Moo". This has now been corrected.

